# Prevalence and types of lower limb conditions in Nepal

**DOI:** 10.1101/2025.01.09.25320193

**Authors:** L Gates, A Channon, A Dickinson, BD Pandey, A Vaidya, B Vaidya, Y Niraula, R Baskota, S Nakarmi, C Metcalf, K Ward, A Silman, A Woolf, MC Puri

## Abstract

**Background:** Understanding the burden of lower limb conditions (LLCs) is essential for effective health planning, yet data in low-resource settings like Nepal is scarce. LLCs, encompassing diverse issues such as pain, injuries and amputations, can profoundly impact mobility, well-being, and livelihoods. With limited healthcare access and growing non-communicable diseases, the scale and impact of LLCs in Nepal remain unknown. This study estimates the prevalence and distribution of person-reported LLCs, exploring their effects on daily living and regional disparities.

**Methods:** To estimate the prevalence of LLCs a household survey was conducted in three selected districts of Nepal representing different ecological zones; Dolakha (Mountain), Lamjung (Hill) and Dang (Terai/Plan). Follow-up questions to 500 individuals with LLCs explored the conditions further, including their distribution by cause and by important characteristics at the individual and ecological zone levels.

**Results:** A total of 2,525 households were surveyed and screened for LLCs. Six hundred and seventy one (26%) of households reported that there was at least one person with an LLC, and at the individual level 11.2% of adults reported an LLC. Sixty-five percent of LLC sufferers were women. Pain and discomfort was the most common problem (reported in 97% of those with an LLC), followed by injury/trauma (19%), deformity (7.2%), wounds (1.4%) and amputation (0.8%). We present the regions of the lower limb most affected, causes of conditions and the extent to which each condition affected participants’ ability to carry out activities of daily living.

**Conclusion:** Our study shows a high prevalence of LLCs across diverse regions of Nepal, with pain and discomfort most frequently reported, and injuries and trauma common in specific areas. LLCs impair daily activities and employment, highlighting the need for tailored interventions and assistive technologies. Findings pave the way for larger-scale studies and scalable, cost-effective solutions.

## Introduction

Understanding the burden and pattern of health conditions is vital for health systems in order to plan and offer the services needed for the population. However, in many lower resource settings, this information is only known through the Global Burden of Disease (GBD) studies, which often use modelled estimates in broad condition groups (1). Further, many countries’ health systems are focused on the communicable disease burden, with certain non-communicable diseases also given high priority. A focus on specific conditions that may have profound consequences on the lived experiences of individuals is often missed, including those affecting the lower limb. These lower limb conditions (LLCs) can lead to pain, mobility limitation, disability, and psychological impact while affecting individual well-being, the household, the wider community, and society. The burden and pattern of LLCs in the population is not well known.

LLCs encompass a wide range of issues affecting the hip, thigh, knee, calf, ankle or foot, and may include congenital abnormalities, neurological, vascular, musculoskeletal pathologies, injuries, or amputation (2-4). There is global evidence for the prevalence of conditions such as rheumatoid arthritis (RA) (5), osteoarthritis (OA) (6), low-back pain (LBP) (7), and gout (8), all of which are recognised broadly as musculoskeletal conditions in the GBD Study (9). Although musculoskeletal conditions comprise about 150 discrete conditions, the GBD specifically reports on health estimates for hip and knee OA, RA, back and neck pain, gout, and a group of ‘other’ musculoskeletal conditions, which refers to a range of conditions that vary across epidemiologic studies (10). Most require diagnosis via sophisticated assessment and clinical expertise, and thus are often underreported in Low- and Middle-Income Countries (LMICs). Chronic pain is a common symptom and thus indicator for many of these conditions (11).

Given the social and economic variation in many LMICs, it is important to capture the impact of LLCs across a range of communities and diverse geographical regions. Once this is known, effective, accessible, acceptable, and low cost strategies for treatment can be developed that will reduce the burden and impact of LLCs. Nepal is an LMIC with many different terrains and over one-third of the population living in rural areas (12) with limited access to good quality healthcare specifically related to LLC treatment and care. Additionally, Nepal faces an increasing non-communicable disease burden(13), including diabetes, which can lead to LLCs, coupled with poor footwear and an increasing number of road traffic accidents (14). The scale of LLCs in Nepal is unknown, alongside the healthcare-seeking behaviours and the coping strategies for those suffering from these with respect to work, accessing treatment, assistive devices, and general healthcare, and economic hardship.

One significant challenge that hampers the estimation and surveillance of LLCs in LMICs is the difficulty of collecting accurate data in a population with limited access to diagnostic modalities, health care records, and health registers. To understand the impact of LLCs it is important to capture all symptoms that may affect daily living and mobility. Thus a holistic self-report approach to LLC criteria may be a more effective way of estimating their scale and impact than using diagnostic tools focused on a specific condition.

The limited available evidence suggests that approximately 2.35 million people in Nepal live with musculoskeletal conditions, with a prevalence of 14.8% and an unmet need of treatment of 60%. Over a third of these cases are estimated to relate to the lower limb (15). A recent cross-sectional community-based study, which adopted the control of rheumatic diseases (COPCORD) tool, estimated the prevalence of musculoskeletal pain/swelling (past and current) in a single rural area was 26.81% (95% CI 24.9–28.7) (16). As LLCs encompass a broader range of conditions, this suggests even more of the population may suffer from an LLC in some form. We do not know the distribution of people with these conditions across different areas in Nepal, nor the impact of these conditions on Nepalese society. Currently, there is no specific institution within the disability management section of the Ministry of Health and Population in Nepal that focuses on the surveillance and care of LLCs.

This study aims to estimate the prevalence of a broad range of person-reported LLCs, exploring their distribution by type of condition and by important characteristics at the person and ecological regional levels, and the effect they may have on activities of daily living.

## Methods

### Study Population

To obtain prevalence estimates, a total of 2,525 households were screened for LLCs. A total of 500 men and women with LLCs were interviewed from three districts; Dang (n=200), Dolakha (n=100), and Lamjung (n=200).

### Sampling

A three-staged cluster sampling technique was used to select men and women with LLCs in households in three selected districts of Nepal, based on its three ecological zones; Mountain (Dolakha), Hill (Lamjung)and Terrai/Plain (Dang). In each district, a ward of a municipality was considered as a cluster and served as the Primary Sampling Unit (PSU). From the list of clusters, four clusters (two from urban and two from rural) in each district were chosen following a systematic probability proportional to size to (PPS) method. A total of ten clusters (four from the hill and plain districts and two from the mountain district) were selected. The sample size in Dolakha was agreed to be smaller due to the greater access issues to the district and the larger distances between villages. Household screening was conducted on 2,525 households across three districts (Dolkha, Lamjung and Dang) of Nepal. Of these first five hundred men and women to report an LLC were selected for interview.

### Inclusion Criteria

Each household was screened for eligible participants based on the inclusion criteria of being above the age of 18 years old, with a self-reported lower limb condition defined as a current (new or longstanding) problem with their leg or foot.

### Questionnaire Design and Surveys

Survey questions sought to estimate the number and type of self-reported LLCs, along with treatment seeking and the impact LLCs have on daily living. Five lower limb condition categories were investigated including pain and/or discomfort, deformity, amputation, wounds, and injury and trauma. The lower limb was defined as the hip, thigh, knee, calf, ankle or foot.

The definition of self-reported conditions were adapted from the Global Alliance for Musculoskeletal Health survey module (17), Community Oriented Program for Control of Rheumatic Diseases (COPCORD) tools (18) the Washington Group questions (19) and the WHO Disability Assessment Schedule (20) finalised in consultation with advisory committee members from for the purpose of this study (Appendix 1). In summary, pain and/or discomfort was defined as defined as any pain or discomfort affecting muscles or joints in the lower limb in the past month, reported by the participants by indicating from 26 reference pain locations provided on a lower body manikin (Appendix 2). ‘Amputation’ was based on any previous amputation at the leg or foot. ‘Lower limb deformity’ was either congenital or acquired. ‘Injury or trauma’ was any that had been suffered that had left a lasting effect on the lower limb, and ‘wounds’ was an open sore on the leg or foot in the past month that had taken over 2 weeks to heal. Participants could report more than one condition and the location of condition.

Questionnaires were first developed in English and then translated into Nepali. We implemented necessary modifications to the questionnaire before full implementation, after pre-testing it with 12 adult men and women with LLCs (from 41 households) in the outskirts of Kathmandu. Questionnaires were programmed into the data collection software KoboToolbox (Kobo Inc., Toronto, Canada). Data were collected between July-August, 2021. Eleven research assistants were trained by a UK-qualified Podiatrist (LG) in the recognition and understanding of lower limb conditions. They obtained written informed consent and conducted survey interviews in person in a private space in participants’ homes using electronic tablets. Participants who were illiterate provided thumbprints to confirm consent.

### Data analysis

All analysis was completed in Stata version 17.0 (Stata Corp, College Station, Texas, USA). Prior to analysis, data distributions were checked for inconsistencies, outliers, and missing information.

Baseline characteristics are described using means with standard deviations, and frequency counts and percentages. The prevalence of an LLC was calculated based on both the number of households and the number of participants with at least one LLC. The frequency of LLC was stratified by geographical region, type of LLC, and body region.

## Results

A total of 2,525 households were surveyed and screened for individuals with LLCs. Six hundred and seventy-one (26%) households reported that there was at least one person with an LLC, and at the individual level, 11.2% of adults reported an LLC. Table 1 describes this population according to age, sex of the head of households, regions, and presence of LLC.

**Table 1.** Description of Households Screened in the overall survey (N = 2525)

| **Characteristics** |  |
| --- | --- |
| <u>Age of household member yrs, mean (SD)</u> | 32 (21.6) |
| <u>Sex of head of the household, n (%)</u> |  |
| Female | 1529 (60.6) |
| Male | 996 (39.4) |
| <u>District Name, n (%)</u> |  |
| Dang | 1006 (39.8) |
| Dolakha | 500 (19.8) |
| Lamjung | 1019 (40.4) |
| <u>Presence of at least one LLC in household, n (%)</u> |  |
| Present | 671 (26.6) |
| <u>Type of LLC, n (%)</u> |  |
| Pain or discomfort | 486 (19.2) |
| Injury/trauma | 95 (3.8) |
| Deformity | 36 (1.4) |
| Wound | 7 (0.3) |
| Amputation | 4 (0.2) |

Of the 500 participants who answered the LLC questionnaire, the majority were female and had a mean age of 57 years. Table 2 describes this population by district according to important demographic factors. Between 8 and 19% of LLC sufferers self-reported having osteoarthritis in at least one lower limb joint, however limited access to diagnostic assessment means this is likely not to be an accurate estimate of radiological osteoarthritis. In Dolakha (Mountain environment) one in every five reported having a fracture that affected their current mobility. Self-reported osteoarthritis, diabetes, and neurological conditions were highest in Dang (Terai/Plain).

**Table 2.** Description of individuals reporting an LLC (N = 500)

|  | **Dang**<br>**(Terai)**<br>**(n=200)** | **Dolakha**<br>**(Mountain)**<br>**(n=100)** | **Lamjung**<br>**(Hill)**<br>**(n=200)** |
| --- | --- | --- | --- |
| <u>Age yrs, mean (SD)</u> | 55.8 (15.9) | 56.2 (13.2) | 58.1 (14.3) |
| <u>Sex of head of the household n (%)</u> |  |  |  |
| Female | 128 (64.0) | 66 (66.0) | 132 (66.0) |
| Male | 72 (36.0) | 34 (34.0) | 68 (34.0) |
| <u>Medical conditions n (% yes)</u> |  |  |  |
| Osteoarthritis | 38 (19.0) | 8 (8.0) | 30 (15.0) |
| Fractures | 7 (3.5) | 14 (14.0) | 2 (1.0) |
| Diabetes | 22 (11.0) | 8 (8.0) | 20 (10.0) |
| Neurological conditions* | 12 (6.0) | 6 (6.0) | 11 (5.5) |

\*Including stroke, Parkinson's, cerebral palsy, spinal cord injury, multiple sclerosis and other neurological disease

In total 628 counts of five lower limb condition categories were reported (Table *3*), indicating differences by district, sex, and age. Pain and discomfort was the most common problem, with 97% reporting lower limb pain or discomfort in the past month. Pain and discomfort and injury and trauma were more commonly reported in females, with over two-thirds of females reporting each of these. Nineteen percent reported having a lower limb deformity, and one percent reported a wound or amputation.

**Table 3.** Percentage of individuals reporting different conditions by background characteristics.

|  | **Pain or Discomfort**<br>**n (%)** | **Injury/Trauma**<br>**n (%)** | **Deformity n (%)** | **Wound**<br>**n (%)** | **Amputation**<br>**n (%)** |
| --- | --- | --- | --- | --- | --- |
| *Sex* |  |  |  |  |  |
| Male | 162 (33.3) | 29 (30.5) | 21 (58.3) | 6 (85.7) | 4 (100.0) |
| Female | 324 (66.7) | 66 (69.5) | 15 (41.7) | 1 (14.3) | 0 (0.0) |
| *Age* |  |  |  |  |  |
| 18-39 | 63 (13.0) | 14 (14.7) | 11 (30.6) | 1 (14.3) | 2 (50.0) |
| 40-54 | 138 (28.4) | 33 (34.7) | 6 (16.7) | 1 (14.3) | 1 (25.0) |
| 55-69 | 182 (37.5) | 31 (32.6) | 10 (27.8) | 3 (42.9) | 1 (25.0) |
| 70+ | 103 (21.2) | 17 (17.9) | 9 (25.0) | 2 (28.6) | 0 (0.0) |

The proportion of people living with an LLC, was generally highest in the region of Dolakha, except for wounds which were observed most often in Dang (Table 4). The distribution of those with pain/discomfort and deformity was reasonably similar across the regions. The proportion of people living with injury /trauma was significantly greater in Dolakha compared to Dang and Lamjung (P<0.001).

**Table 4.** Proportion of individuals with each type of LLC (compared to without) by region.

|  | **Dang**<br>**(n=200)** | **Dolakha**<br>**(n=100)** | **Lamjung**<br>**(n=200)** |  |
| --- | --- | --- | --- | --- |
| Pain or Discomfort, n (% yes) | 192 (96.0) | 99 (99.0) | 195 (97.5) | p=0.314 |
| Injury/Trauma, n (% yes) | 48 (24.0) | 26 (26.0) | 21 (10.5) | p<0.001 |
| Deformity, n (% yes) | 14 (7.0) | 8 (8.0) | 14 (7.0) | p=0.942 |
| Wound, n (% yes) | 6 (3.0) | 0 (0.0) | 1 (0.5) | - |
| Amputation, n (% yes) | 0 (0.0) | 3 (3.0) | 1 (0.5) | - |

\*differences tested using chi-squared ( $\chi^2$ ). Differences in the proportion of wounds and amputations between countries were not tested due to low counts

Of 500 people with an LLC, 486 (97%) reported pain or discomfort in lower limb muscles or joints in the past month. These were grouped into either hip, knee, foot/ankle, thigh, or lower leg, and the distribution of these is shown in Figure 1. Over 70% reported pain in the knee, followed by 48% at the foot/ankle. A high proportion reported symptoms in more than one location (Figure 2), with three people reporting pain in all twenty-six lower limb regions.

**Figure 1:**
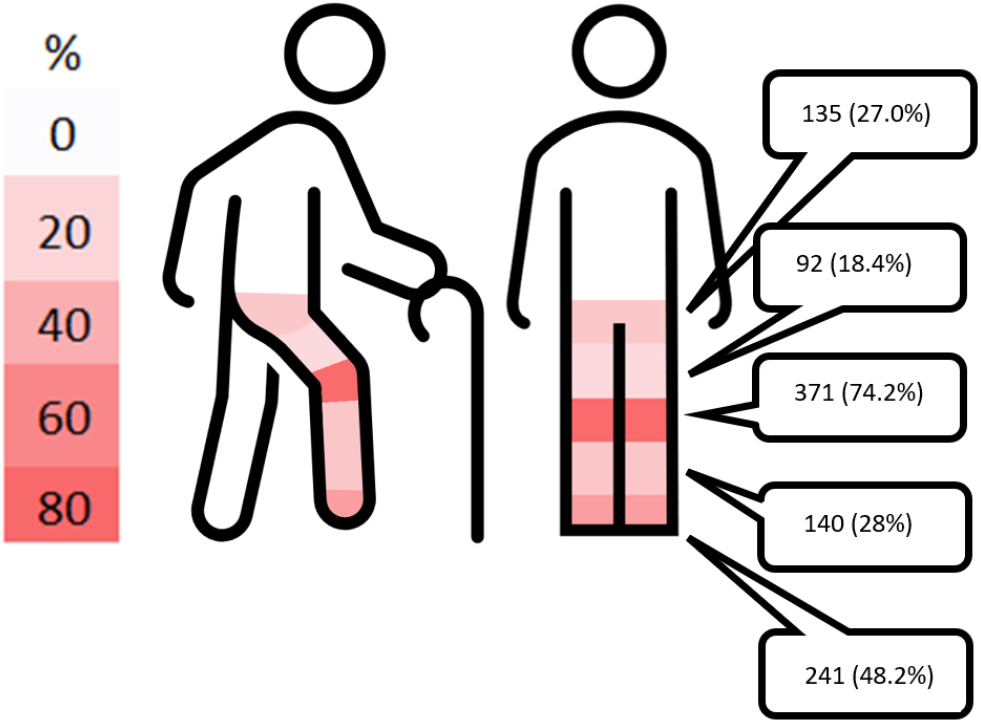
Prevalence of self-reported pain or discomfort in the hip, thigh, knee, calf, ankle or foot, reported as n (%)

**Figure 2.**
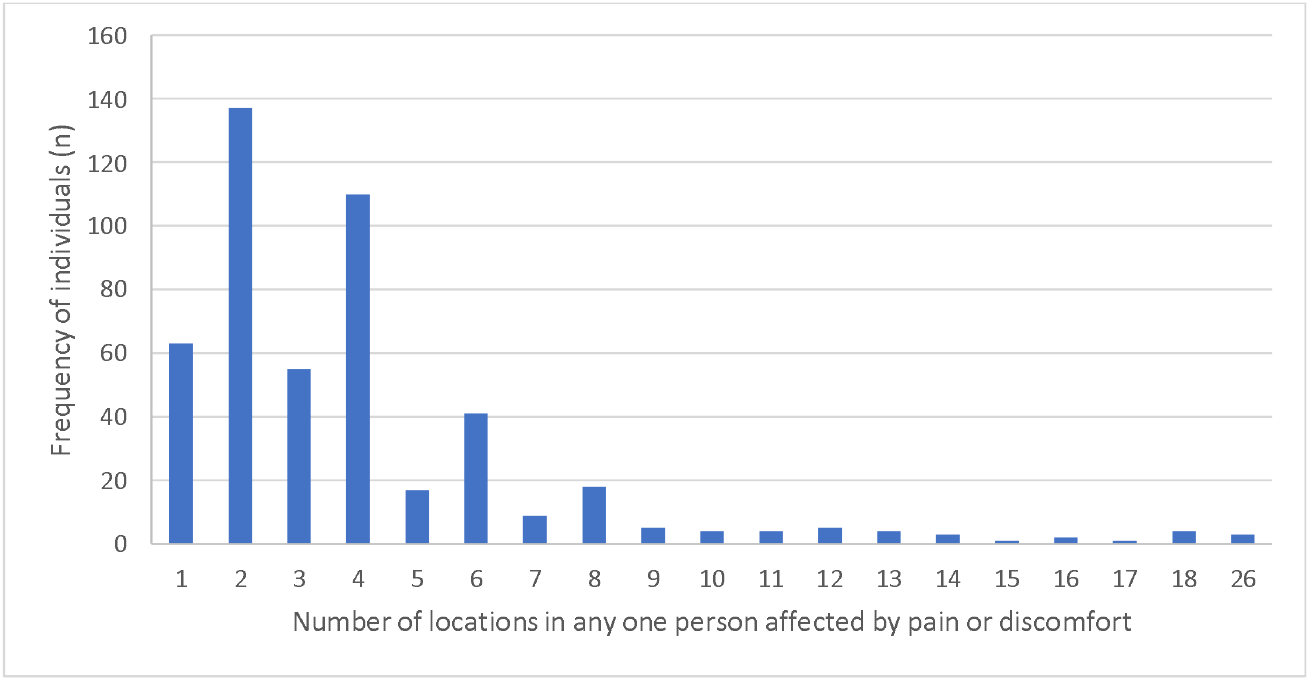
Frequency of individuals reporting one or more locations for lower limb pain or discomfort

Of the 36 respondents with a lower limb deformity, the causes of these deformities included congenital (31%), trauma (22%), disease (19%) and other causes (28%). Of those who had sustained lower limb injury/trauma (n=95), 66% were from a fall, 14% due to a vehicle accident, 13% from an agricultural accident and 7% from other mechanism. Additionally 16 people (3%) reported ever sustaining a hip fracture. The cause of wounds (n=7) included leprosy (14%) and other causes (86%). Reasons for lower limb amputation (n=4) included trauma, burns, cancer and nerve related problems.

Figure 3 shows the extent to which each condition affected participants’ ability to carry out activities of daily living. All participants with amputation reported that their condition severely or extremely affected their ability to carry out activities of daily living (ADLs). Likewise, 82% percent of those with a musculoskeletal condition, 71% with a wound, 69% with injury/trauma, and 55% with deformity reported their condition severely or extremely affected their ability to carry out ADLs. Only in cases of deformity (5.5%) and injury/trauma (10%) did any participants report their condition did not affect ADLs.

**Figure 3.**
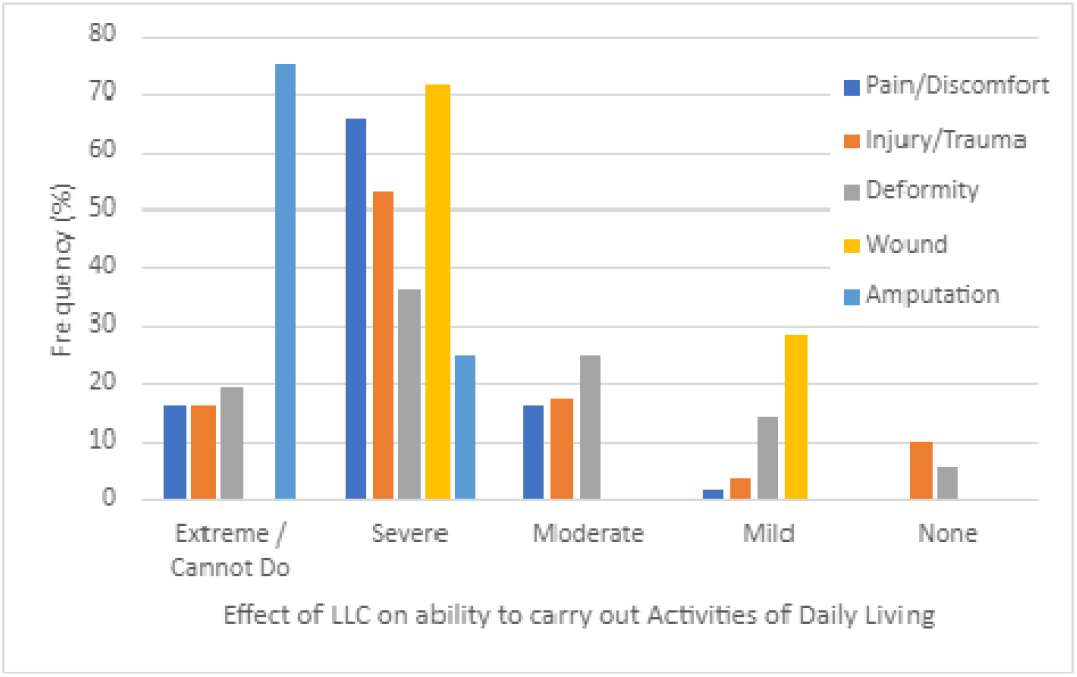
The frequency of severity for which an LLC affects participants’ ability to carry out activities

Individuals with LLCs were asked about their current occupation. Ten percent (n=50) stated that they were unemployed, with 96% (n=48) of these stating that this was due to a health condition or disability. However, a majority of these unemployed individuals were aged over 60 (88%, n=44) so are also near or older than the official retirement age (with only a further 7 individuals reporting that they were retired). A further 41.4% (n=207) reported that their occupation was ‘household chores’, with most (90%) doing this in their own home rather than in a different home. This was at all ages, potentially indicating the limited ability of LLC sufferers to obtain an occupation. Over 70% of those who conducted HOUSEHOLD chores stated that they had given up their work due to their LLC, indicating that the chores were not a choice of occupation for many.

## Discussion

This is the first known study to investigate the distribution and impact of a broad range of LLCs in Nepal. Of all households screened across three regions with different ecologies, 26% reported at least one person with an LLC, and 11% of all adults screened reported having an LLC. Pain and discomfort were the most prevalent, with 19% of all surveyed participants reporting symptoms.

Our findings reveal a relatively similar distribution of pain/discomfort and deformities across the three regions of Nepal. However, a higher proportion of injuries and trauma was reported in Dolakha and Dang, accounting for 26% and 24% respectively, compared to 11% in Lamjung. Furthermore, wounds were predominantly reported in Dang, while amputations were more prevalent in both Dang and Dolakha, although the number of these conditions were small. Different factors may contribute to the higher frequency of injury and trauma in the Dang and Dolakha regions. Dang is likely to experience an increased likelihood of workplace accidents or injuries, being an area with a higher proportion of people involved in agricultural work, labour-intensive industries, and activities. Dolakha on the other hand is a mountain district with more rugged terrain, where geographical features and topography may heighten the risks of accidents and injuries. Additionally, socio-economic factors such as infrastructure development, road conditions, and access to healthcare services differ between regions. Cultural and behavioural factors may also be related to risk-taking behaviours, safety practices, and awareness of injury prevention measures might also vary between regions, influencing the reporting rates of injuries and trauma.

These differences between regions emphasize the varying patterns of health challenges faced within Nepal, emphasizing the importance of tailored healthcare interventions to address specific needs effectively. As per the recent Department of Health Services of Nepal annual report (21), progress is being made across disability-inclusive health, rehabilitation, assistive technology, and injury prevention. For example, Nepal has seen the piloting of a disability management and rehabilitation training package for primary health care providers, plus the development of the national standard on assistive technology and priority assistive product list. Furthermore, the Systematic Assessment of Rehabilitation Situation report (STARS) (22) was finalized and the Rapid Assistive Technology Assessment (rATA) (23, 24) was conducted in coordination with the National Health Research Center. Preliminary data has been collected to evaluate the rehabilitation workforce using WHO-standardised tools. Our results highlight the impact of various LLCs on individuals’ ability to perform activities of daily living and undertake their chosen occupation in Nepal. Notably, all participants with amputation reported that their condition severely or extremely affected their daily activities, indicating the significant functional limitations faced by this subgroup. Furthermore, the high percentages of individuals reporting severe or extreme limitations in carrying out activities of daily living across other LLCs are notable. These findings highlight the broad-ranging impact of LLCs on individuals’ functional abilities and quality of life, regardless of the specific nature of their condition.

Among those with a LLC, 10% were not currently employed, with 96% attributing this status to a health condition or disability. Additionally, over three-quarters reported that their LLC influenced their choice of occupation. These findings highlight the potential impact of LLCs on employment status and career decisions, emphasizing the need for tailored support and accommodations to facilitate meaningful participation in the workforce. Musculoskeletal disorders are associated with high costs to employers and employees including absenteeism, lost productivity, increased health care expenses, disability, and worker’s compensation costs (1, 25, 26). In low-resource countries, where access to sickness and disability allowance is limited or inequitable, this places a much higher burden on those affected by LLCs and their families.

Our findings report the prevalence and distribution of pain or discomfort among individuals with LLCs. A substantial majority, 97% of the participants with an LLC and 19.2% overall (based on the household sample), reported experiencing pain or discomfort in lower limb muscles or joints within the past month. This is comparable to a recent rural survey which reported a 27% prevalence of past and current musculoskeletal pain/swelling (16). This high prevalence highlights the significant burden of musculoskeletal issues in this population. Musculoskeletal pain, especially joint and back pain, is the most common type of chronic pain and is the main contributor to disability worldwide (27). According to the WHO, 20–33% of the world’s population has some form of chronic musculoskeletal pain, translating to 1.75 billion people globally. Data from the Global Burden of Disease study showed the prevalence of other musculoskeletal disorders including a wide range of joint, ligament, tendon, or muscle problems that cause regional or generalised pain was 8.4% (95% uncertainty interval (UI) 8.1% to 8.6%) (10). Global evidence for more specific and commonly investigated conditions suggests a prevalence of between 3.8% and 22.9% (28, 29) for knee osteoarthritis and between 0.85% and 8.55% for hip osteoarthritis (28, 30). This range in estimates provides some insight into the difficulty of comparing even specific conditions, in part due to differences in pathological definitions.

The only other known study to capture lower limb-specific data in Nepal suggests that the country has approximately a 14.8% prevalence of people living with musculoskeletal conditions. Over a third of these were related to the lower limb (15). However, making comparisons is difficult due to differences in condition definitions and anatomical regions used in various studies.

The categorization of pain into specific regions such as the hip, knee, foot/ankle, thigh, or lower leg provides valuable insights into the localization of symptoms. Notably in this study, the knee was the most commonly affected area, with over 70% of individuals reporting knee pain, followed by 48% experiencing discomfort in the foot/ankle. This distribution suggests that certain anatomical sites within the lower limb are more susceptible to pain among individuals with lower limb conditions. Evidence from high-income countries also suggests knee is the most frequently affected of all joints (31, 32).

Our prevalence estimates are difficult to compare to those of other countries given there is little research into LLCs defined as broadly as in this study. These findings cannot be generalised to the entire country due to small sample size. However, utilising a holistic approach to defining LLCs has enabled us to gather information on a significant number of people, including those in very remote areas. We appreciate the limitation of self-reported conditions and the fact this may bring with it an over-estimation of conditions. However, this was judged a necessary compromise given that there is limited access to diagnostic health care in many regions of Nepal. Social desirability bias may also be present, where respondents may have provided answers to questions in such a way as to present themselves in socially acceptable terms.

## Conclusions

Due to the absence of quantifiable evidence for the impact of LLCs, especially in LMICs, the opportunities to develop and influence strategies for their management have been limited. A number of LLCs are potentially modifiable to improve the health and lives of sufferers. However, without knowledge of their epidemiology, impact, and subsequent need for treatment we are limited in our ability to expand translational research and to develop effective services. The generation of population data to estimate the impact of LLCs now leverages opportunities to target society-driven solutions and interventions in Nepal.

This study provides preliminary information about the scale of LLCs in Nepal. The findings raise the profile of this neglected widespread area and have set the context where a larger-scale investigation can be conducted, which will also provide simple and effective intervention strategies that can be implemented at scale to mitigate the effects of these conditions. The value of investment in mobility assistive technologies is already clear, reported by ATscale at 9:1 for devices including prostheses and wheelchairs (33).

Based on our findings, we provide the Nepali government and organisations with options to consider while developing and implementing effective and targeted policies to drive Nepal’s efforts in addressing the UN Sustainable Development Goals for Good Health and Wellbeing (SDG3) and reduced inequalities (SDG10). These will inform specific strategies moving forward in the government agenda for NCDs and for the health care system reform towards universal health coverage. These considerations are:

- **Support for individuals with LLCs who want and are able to work**. Ensuring there is high-level support for people with disabilities to continue with their work, or bridging loans to ensure that individuals can recommence work after recovery, will benefit individuals and the economy.
- **Increase the supply of trained professionals in rehabilitation and lower limb medicine**. Building capacity of trained staff, for example, Podiatrists, Prosthetists and Orthotists, and Occupational Therapists, should improve the ability to provide medical intervention, rehabilitative management, devices, and aids, as well as advice on adapting the built environment for people with impaired mobility.

### Additional considerations

- **Create equitable access to services and devices**. Equitable access may be enabled by i) signposting to appropriate professionals in health care facilities, and ii) Investing in mobility devices that are fit for purpose and accessible to all.
- **Design an appropriate built environment**. Where possible, accessibility for people with mobility issues should be included at the design stage for planned building projects and community development.

## Acknowledgments

We thank the field staff of the Center for Research on Environment Health and Population Activities (CREHPA) who diligently collected data from all three study areas in Nepal.

## Funding Statement

We gratefully acknowledge support from the Economic and Social Research Council (ESRC) as part of the Global Challenges Research Fund (GCRF) scheme (University of Southampton internal grant).

### Ethical Approval and Informed Consent Statements

Ethical approval was obtained from the University of Southampton (ERGO Reg No 62291) and Nepal Health Research Council (Reg No 51/2021 P). All of the participants agreed to voluntarily join the study and subsequently sign the informed consent forms.

### CRediT authorship contribution statement

Gates L: Conceptualization; Formal analysis, Funding acquisition, Investigation, Methodology, Project Administration, Writing - original draft, Writing - review & editing. Channon A: Conceptualization; Formal analysis, Funding acquisition, Investigation, Methodology, Project Administration, Writing - original draft, Writing - review & editing. Dickinson A: Formal analysis, Writing - original draft, Writing - review & editing. Pandey B: Methodology, Validation, Writing - review & editing. Vaidya A: Methodology, Writing - review & editing. Vaidya B: Methodology, Writing - review & editing. Niraula YR: Methodology, Writing - review & editing. Baskota R: Writing - review & editing. Nakarmi S: Writing - review & editing. Metcalf C: Methodology, Writing - review & editing. Ward K: Methodology, Writing - review & editing. Silman A: Methodology, Writing - review & editing. Woolf A: Methodology, Writing - review & editing. Puri M: Conceptualization; Data Curation, Formal analysis, Funding acquisition, Investigation, Methodology, Project Administration, Supervision, Writing - original draft, Writing - review & editing.

## Data Availability Statement

The data that support the findings of this study are available on request from the corresponding author.

## Appendix A

**Table A.1.** Definitions for each condition.

| Condition | Definition | Recall period |
| --- | --- | --- |
| Pain and/or discomfort (musculoskeletal condition) | Any pain or discomfort affecting your muscles or joints in your lower limb (hip, thigh, knee, calf, ankle or foot)<br>→ Followed by indicating from 26 reference pain locations provided on a lower body manikin (appendix 3) | Past month |
| Amputation | Previous amputation at the leg or foot<br>→ Followed by a choice of multiple locations to the indicated region of amputation | Ever |
| Deformity | Any deformity you may have at your leg or foot, that you were born with or acquired at any age<br>→ Followed by indicating from 26 reference pain locations provided on a lower body manikin | Ever |
| Injury or trauma | An injury or trauma to your lower limb that has left a lasting effect on your leg<br>→ Followed by indicating from 26 reference pain locations provided on a lower body manikin | Past month |
| Wound | An open sore on your leg or foot that has taken over 2 weeks to heal – (This does not include a blister, verruca, bruise or rash)<br>→ Followed by a choice of multiple locations to indicate region of amputation | Past month |
| Other conditions | Any altered sensation (numbness/tingling, muscle weakness) in your legs or feet<br>→ Followed by a choice of multiple locations to indicate region of amputation | Past month |

## Appendix B

**Figure A.1.**
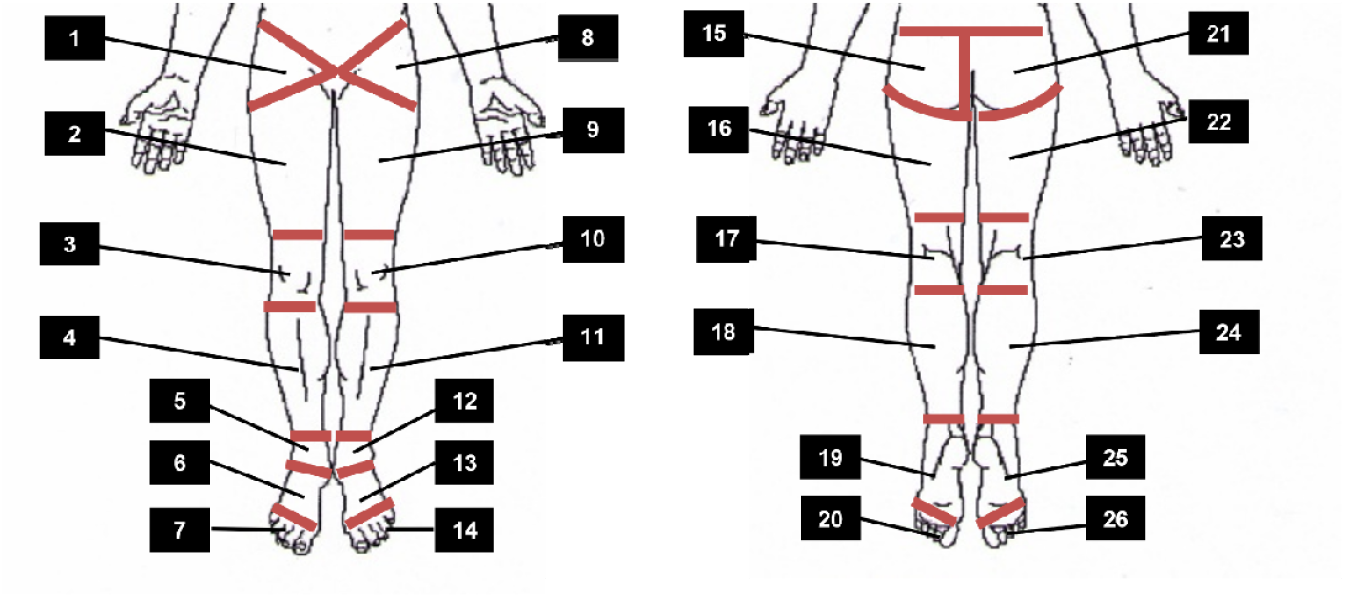
Regions to report lower limb pain/discomfort on mannequin

